# A Web Application for Exploring Distribution in Academic Publications Across Geography and Institutions in India

**DOI:** 10.64898/2026.03.18.26348755

**Authors:** Yiren Hou, Evan Cohen, Jackson Higginbottom, Lillian Rountree, Yi Ren, Brian Wahl, Kate Nyhan, Bhramar Mukherjee

**Affiliations:** Department of Biostatistics, Yale School of Public Health, New Haven, CT 06510, USA; The Computer Science and Engineering Division, University of Michigan, Ann Arbor, MI 48109, USA; Department of Epidemiology of Microbial Diseases, Yale School of Public Health, New Haven, CT 06510, USA; Harvey Cushing/John Hay Whitney Medical Library, New Haven, CT 06510, USA; Department of Chronic Disease Epidemiology, Yale School of Public Health, New Haven, CT 06510, USA; Department of Statistics and Data Science, Yale University, New Haven, CT 06511, USA

**Keywords:** Publication trends, Data visualization, Human Development Index

## Abstract

India’s national research capacity and infrastructure are unevenly distributed across states and union territories (UTs), contributing to geographic variation in academic publication output. We developed **Indiapub**, an open-access web application that quantitatively enumerates and visually displays geographic and temporal publication patterns for research products with at least one author affiliated with an Indian institution, using OpenAlex data. The app is designed for ease of use, with automated data retrieval, cleaning, and aggregation. **Indiapub** allows users to filter publications by topic, publication year range, author position, publication type, minimum citation count, state/UT, and population size of the state/UT where the author institution is located. The app also provides downloadable tables and ranked institution lists by publication count. Its interactive dashboard includes five modules: (i) a map of publication distribution, (ii) time trend plots for nation and state/UT, (iii) publication-share versus population-share plots highlighting over- and underrepresentation, (iv) stacked bar charts of state/UT contributions over time with population benchmarks, and (v) bubble plots relating the Human Development Index to publication volume over time. This tool may support resource prioritization and identification of institutional strengths for trainees, researchers, higher education administrators, and policymakers. To illustrate its utility, we present sample findings derived from the app. For publications across all topics from 2014 to 2025, the largest research participation footprints were observed in Tamil Nadu, Maharashtra, Delhi, Uttar Pradesh, and Karnataka. Tamil Nadu and Delhi were home to three of the highest-publishing institutions nationally: Vellore Institute of Technology, All India Institute of Medical Sciences, and Indian Institute of Technology Delhi. We also examined six curated case studies of broad scientific interest: electronic health records (EHR), genome-wide association studies (GWAS), artificial intelligence (AI), development economics, environmental science, and COVID-19. Findings from these case studies revealed over- and underrepresentation in publication output across states and UTs. For example, in EHR publications among high-population states, Tamil Nadu’s publication share exceeded its population share by 31.3 percentage points (pp), whereas Bihar’s was 12.8 pp lower. Our tool offers insights into India’s research landscape across states and UTs with easy-to-digest visuals. Such interactive tools have the potential to serve as a starting point for fostering a more inclusive research ecosystem supporting targeted research policy and planning.

## Introduction

Global scientific output has expanded rapidly in the last decade. In 2022, six countries—China, the United States, India, Germany, the United Kingdom, and Japan—each produced more than 100,000 Scopus-indexed publications in science and engineering, including peer-reviewed journal articles and conference papers [1]. India is a major contributor to this growth, yet an aggregated national summary can mask large within-country differences. In a country that is so diverse in its socioeconomic, linguistic, and institutional landscape, variations in research capacity and infrastructure are inevitable, but the granular details of how these variations impact research outcomes are unknown and difficult to quantify. Understanding variations in research participation through publication output over time at a subnational level in India is a strong piece of data for developing equitable research and educational policy, finding institutions of excellence within specific regions and within specific research domains, and planning investments to bridge identified gaps.

Several tools can help analyze heterogeneity in research output. Bibliometric platforms such as CiteSpace II [2,3] and VOSviewer [4] help researchers map the research landscape by retrieving records from large databases (e.g., PubMed, Web of Science) and generating visual maps that reveal the growth of research topics and the connections among publications, authors, and keywords. However, these tools are primarily geared toward technical users and are not designed to visualize subnational publication trends over time. In addition, these platforms do not seamlessly link to external data sources, limiting the exploration of how research productivity relates to regional population size or other metrics of human development, such as the Human Development Index (HDI) [5]. While regional visualizations exist on discovery platforms, such as Scinapse.io (Pluto, Inc.) [6,7], Dimensions.ai [8], and Lens.org [9], access can be restricted (e.g., subscription requirements), and state- and territory-level publication trajectories across time are often limited. Regarding India in particular, India-focused dashboards like Harvard’s India Policy Insights [10] provide accessible spatiotemporal views for health, education, and development indicators, while Data for India [11] is a public platform that analyzes population, health, economy, living conditions, work, and measurement across the nation. However, neither Harvard’s India Policy Insights nor Data for India includes research publication data or offers tools to track or analyze publication output at the subnational level, leaving policymakers, funders, and institutional leaders without an India-specific tool to examine publication patterns across states and union territories (UTs).

To fill this gap, we developed an open-access web application (**Indiapub**, accessed either via https://indiapub.org or https://bharatresearch.org) that quantitatively enumerates geographic and temporal patterns of academic publication output in India at the national, state-, and territory-level using publications with at least one author affiliated with an Indian institution. We intend to maintain the **Indiapub** web application and its public access through at least December 2027. This application is designed for ease of use, in which data retrieval, cleaning, and aggregation are automated. Users can interactively filter publications by topic of interest, year range, author position, publication type, minimum number of citations, state and UT, and population groups (high/medium/low). Beyond simple publication counts, the application provides five visualizations: (i) a map that shows geographic distribution of publications; (ii) a time trend plot that shows temporal distribution of publications for national totals and for each state/UT; (iii) a plot of publication-share versus population-share by state and UT plot that highlights over and underrepresentation; (iv) a stacked bar plot showing relative publication contribution of state/UT over time with reference to relative population benchmark; and (v) a bubble plot relating state/UT-level HDI to publication output over time. These analyses are not supported in standard publication databases.

To demonstrate the application’s capabilities and information provided, we present a sample analysis. We first describe the overall publication distribution from 2014 to 2025 for any topics. Then, we provide six illustrative case studies on topics of broad academic interest: electronic health records (EHR), genome-wide association studies (GWAS), artificial intelligence (AI), development economics, environmental science, and COVID-19, based on the publication activity in India from 2014 to 2025. Using the application’s core functions, we identify variations across domains and across states/UTs. A state/UT’s publication share often diverges from its population share, inducing over and underrepresentation. We identify such variations by comparing the publication share versus population share of states/UTs across population groups. We also relate publication patterns to state/UT-level HDI values over the period 2014 to 2025. The web application benefits multiple user communities, including scholars seeking to analyze geographic publication patterns, students and emerging academics exploring regional research opportunities, and higher education administrators, funders and policymakers aiming to identify underrepresented regions for building research capacity in India. By providing transparent metrics and an intuitive interface, our application serves as a practical starting point for evidence-informed planning, capacity building, and cross-regional collaboration in India, with a framework that is adaptable to other countries.

## Methods

### Data retrieval and keyword-based searches

Publication data from 2014 to 2025 were obtained from the OpenAlex Application Programming Interface (API) [12], an open access scholarly metadata database. For predefined topics and custom keyword searches, **Indiapub** retrieves publications by applying keyword-based queries to the title and abstract fields in OpenAlex. By contrast, when users select “Any Topics,” the application does not rely solely on a live keyword-based API query; instead, it uses a precomputed India-wide publication dataset, with optional background loading of the full publication-level records. We used the default OpenAlex data version available via the API at the time of extraction (Walden / Data Version 2). The **Indiapub** application includes a set of recommended topics (including but not limited to those used in our case studies): EHR, GWAS, AI, digital health, COVID-19, development economics, international trade, public finance, macroeconomics, game theory, and environmental science. These topics are implemented as keyword-based searches across the title and abstract fields. In addition to these predefined topics, the application allows users to explore all publications from India when selecting “Any Topics”. Users can also construct their own queries through a “Custom Keyword Search” option in the topic filter. All queries were restricted to publications with at least one affiliation with an institution in India.

When selecting “Any Topics,” users choose between two data loading modules. The choice “Snapshot Only” renders plots instantly using pre-aggregated data and provides top institutions list, but it does not provide access to the individual publication table; author position is fixed to “Any author”; and publication type and citation filters are unavailable. The choice “Snapshot + Live API Loading” displays plots immediately from pre-aggregated data while progressively loading the complete publication dataset in the background, which typically takes two to three hours. Once loading completes, all filters become available, plots update to reflect selected filters, and users gain full access to the data table with export functionality.

### Institutional affiliation and geographic mapping

For each publication, we extracted author institutional affiliations using the Research Organization Registry (ROR) identifiers provided by OpenAlex. We matched all RORs to Indian institutions and their corresponding state/UT using metadata from the ROR registry [13]. We define an India-affiliated author as any author with at least one affiliation with an institution in India. Authors with only non-Indian RORs were not considered India-affiliated. If OpenAlex returned no ROR for an affiliation, or the ROR could not be matched to an Indian institution, we did not attribute that author to India or to any state/UT.

### Software characteristics

Users can interactively filter publications by topic of interest, year range, author position, publication type, minimum number of citations, state and UT, and population-size group (high/medium/low). Counts are computed per publication-state/UT pairing: a given publication can add one publication count to multiple states/UTs, but never more than one publication count to any given state/UT. For attribution, we use each author’s first Indian institution (i.e., the first listed affiliation with an institution in India). The interface allows users to choose among four author position counting rules (see Table 1):

- **First author (default for searches other than “Any Topics”):** a state/UT receives one count if the first author’s first Indian institution is located there.
- **Last author:** a state/UT receives one count if the last author’s first Indian institution is located there.
- **First or last author:** a state/UT receives one count if either the first or the last author’s first Indian institution is located there (a publication contributes a maximum of one count to a state/UT under this rule, even if both authors are from the same state/UT). Because first and last authors may be based in different states/UTs, a single publication can contribute one count to two different states/UTs.
- **Any author (default for “Any Topics” search):** a state/UT receives one count for every author whose first Indian institution is located there; thus, a single publication can contribute multiple counts when co-authors are based in different states/UTs. A publication contributes to a maximum of one count under this rule, even if there are multiple authors from the same state/UT. In “Snapshot Only” mode for “Any Topics” search, this rule is fixed and cannot be changed. In “Snapshot + Live API Loading” mode, users may switch to other author position rules after the data loading process completes.

**Table 1.** Indiapub application author position rules and their interpretations.

| **Author Position setting** | **Rules and interpretations** |
| --- | --- |
| **First author (default)** | Counts the state/UT of the first author's first Indian institution. Interprets where the publication's research lead is based and is a good proxy for early-career location. |
| **Last author** | Counts the state/UT of the last author's first Indian institution. Interprets where senior or supervising leadership is based. |
| **First or last author** | Counts the state(s)/UT(s) of the first or last author's first Indian institution. Interprets leadership presence at either end of the author line. |
| **Any author** | Counts the state(s)/UT(s) of every author's first Indian institution. Interprets participation footprint or breadth of collaboration and is more inclusive but less specific about who led the work. |

To allow for meaningful comparisons across states/UTs of varying population sizes, our application classifies Indian states and UTs into three population groups. Users can select population group(s) of interest by selecting any combination of low, medium, or high population groups. A high population group is defined as states/UTs with an estimated population share of over 5.0% of India’s total population. A medium population group is defined as states/UTs with an estimated population share between 1.0% and 5.0% (inclusive) of India’s total population, and a low population group is defined as those with less than 1.0% of the national population. We chose these *a priori* cutoffs to (i) separate a set of very large states whose totals can dominate national patterns (>5.0%), (ii) create a middle band where most states and the National Capital Territory of Delhi (Delhi) lie (1.0%-5.0%), and (iii) distinguish smaller states and UTs (≤1%). This allows our tool to highlight both absolute and relative contributions to publication output. The population estimates were extracted from tables reporting projected total populations of states and UTs from the *Population Projections for India and States 2011-2036* report published by the National Commission on Population under the Ministry of Health and Family Welfare, Government of India [14].

The application includes six key interactive modules that explore publication data:

1. Map of geographic distribution of publications: This is a map that displays state/UT-level publication counts, where states and UTs are shaded with a color gradient. A darker hue represents a greater number of publications. The display metric can be either total publication counts or per 1,000,000 people. By default, the map displays total publication counts.
2. Data table: This is a downloadable table listing publication metadata, including year, publication type, title, citation count, author name, author position, affiliated institution, and state/UT. A list of the top 50 institutions is readily available to download as well. Note that the data table is not available when using “Snapshot Only” mode for “Any Topics” search: instead, only the list of top 50 institutions is available.
3. Time trend plot of publication count: This is a line graph showing the number of publications per year. The plot can show national data or break it down by state/UT. By default, the plot shows national publication trend.
4. Publication versus population share by state and UT: This visualization compares each state/UT’s share of publications with its share of population under the selected states/UTs. For each state/territory, a circle denotes its publication share. We compute each state/UT’s publication share as the count of publications in that state/UT over the selected year range divided by the total publications within the selected states/UTs over the same time range and filters. A square denotes its estimated population share. We compute each state/UT’s population share as the state/UT’s population divided by the total population within the same selected states/UTs at the latest year of the selected time range. Points are colored green when the publication share exceeds the estimated population share (positive percentage point difference). Points are colored orange when the estimated population share exceeds the publication share (negative percentage point difference). The vertical distance between the circle (publication share) and the square (population share) represents the magnitude of over or underrepresentation for that state or UT.
5. Stacked bar plot of state and UT publication percentage by year: For each year, the bar sums to 100% of publications under the selected filters. Each colored segment shows a state/UT’s share of that year’s total publications. A population benchmark can be displayed, either a single bar showing population shares for the selected states/UTs in the final year or the year-by-year population shares across the time range. This view allows users to examine who contributes to what publication share each year and how that compares to population share.
6. Bubble plot of publications versus HDI over time: This is a scatter plot displaying publication counts (represented by bubble size) against the HDI, with year on the x-axis and HDI on the y-axis. The HDI dataset for Indian states and UTs was retrieved from the Global Data Lab [16], which provides estimates of subnational human development from 1990 to 2022. The HDI estimates for 2023 to 2025 were extrapolated based on the average growth rate. Each bubble within the plot represents a state/UT.

## Results

By selecting “Any Topics” and timestamp 2014 to 2025 with no citation threshold, we found that India produced about 4 million publications in this period. In Table 2, we identify the top five states/UTs by their publication counts; describe temporal trends; list states/UTs where the absolute difference between their share of publications and their share of population is greater than ±10.0 percentage points (pp) in their respective population groups; describe how each state/UT’s share of publications changes over time within its population group; report the main contributors by publication count within their respective population groups; summarize the relationship between HDI and publication volume over time; and report top three institutions by their publication counts. We found that Tamil Nadu, Maharashtra, Delhi, Uttar Pradesh, and Karnataka were the leading participants to the overall publication count from 2014 to 2025. Stratified by population group, the main contributors by publication count in their respective groups were Tamil Nadu, Maharashtra, Uttar Pradesh, and West Bengal in high population group, Delhi and Karnataka in medium population group, and Uttarakhand and Chandigarh in low population group.

**Table 2.**
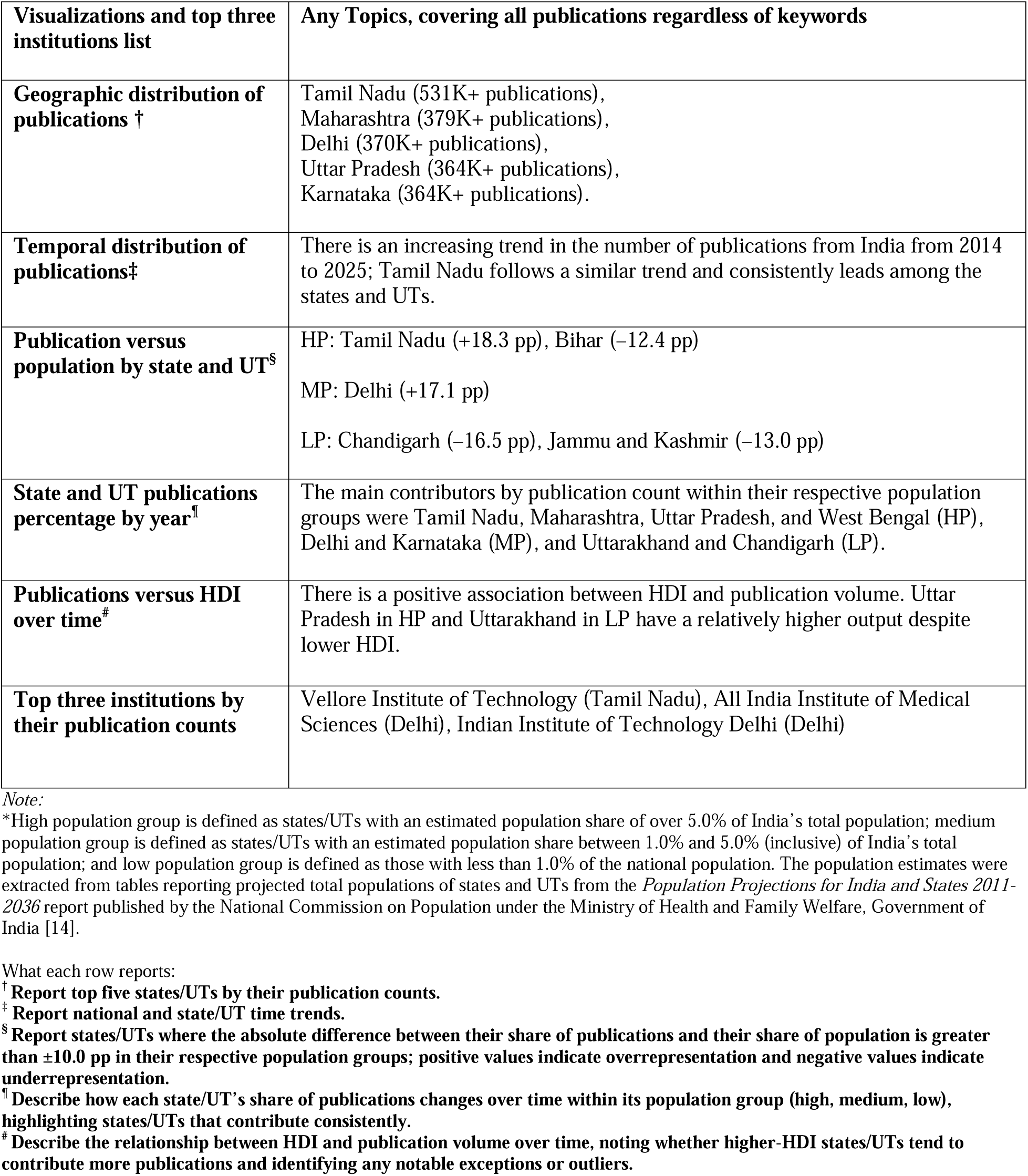
Details of findings from visualizations from the **Indiapub** web application covering any topics from 2014 to 2025, in which “Any author” position is the default in this case. The results derived from the application are as of February 2, 2026. Abbreviations: Thousands (K), High population group (HP), medium population group (MP), low population group (LP), Human Development Index (HDI), National Capital Territory of Delhi (Delhi), percentage points (pp), union territory (UT).

We conducted six case studies on publications from the topics of EHR, GWAS, AI, development economics, environmental science, and COVID-19, covering from 2014 to 2025, inclusive. These areas were chosen to focus on a mix of mature and emerging topics across the health and economics fields. For each case study, we state the top five state/UT contributors; describe the temporal distribution of publications; report over or underrepresentation within high, medium, and low population groups only when a state/UT’s publication share differs from its population share by at least an absolute difference of 10.0 pp; and summarize the relative contributions by state/UT over time and nuances in the relationship between the HDI and publication output. We use the application’s default settings: First author for publication counts, all publication types, and no minimum citations. Details of our findings based on our interactive plots and available data table for each case study are listed in Table 3.

**Table 3.** Details of case study findings from visualizations in the **Indiapub** web application covering the topics of electronic health records (EHR), genome-wide association studies (GWAS), artificial intelligence (AI), development economics, environmental science, and COVID-19 across the period 2014-2025. The results derived from the application are as of March 10, 2026. Abbreviations: High population group (HP), medium population group (MP), low population group (LP), Human Development Index (HDI), National Capital Territory of Delhi (Delhi), percentage points (pp), union territory (UT).

| Case studies<br>(total publications) /<br>Visualization and top<br>three institutions list | Electronic Health Records<br>(3,106 publications) | Genome-wide Association<br>Studies<br>(1,431 publications) | Artificial Intelligence<br>(48,472 publications) | Development Economics<br>(1,750 publications) | Environmental Science<br>(6,432 publications) | COVID-19<br>(55,777 publications) |
| --- | --- | --- | --- | --- | --- | --- |
| **Geographic distribution of publications** † | Tamil Nadu, Maharashtra, Uttar Pradesh, Karnataka, Delhi (Figure 1). | Delhi, Tamil Nadu, Uttar Pradesh, Punjab, Karnataka. | Tamil Nadu, Maharashtra, Uttar Pradesh, Karnataka, Delhi. | Delhi, Uttar Pradesh, Maharashtra, Tamil Nadu, West Bengal. | Tamil Nadu, Uttar Pradesh, Delhi, Maharashtra, Karnataka. | Delhi, Maharashtra, Tamil Nadu, Uttar Pradesh, Karnataka. |
| **Temporal distribution of publications**‡ | Exponential growth in national publication count from 2014 to 2025; Tamil Nadu follows a similar trend and consistently leads among the states and UTs. | Gradual national buildup from 2014 to 2025; Delhi follows a similar trend and consistently leads. | Exponential growth in national publication count from 2014 to 2025 (Figure 2A); Tamil Nadu follows a similar trend and leads among states/UTs (Figure 2B). | Gradual growth to 2021, sharp peak in 2023, followed by an increase of publications from 2024 to 2025; no single state/UT dominates over time. | Exponential growth in national publication count from 2014 to 2025; no single state/UT dominates over time. | Sudden peak in 2021, followed by a rapid decline from 2022 to 2025. Delhi follows a similar trend and leads. |
| **Publication versus population by state and UT**<sup>§</sup> | HP: Tamil Nadu (+31.3 pp); Bihar (–12.8 pp)<br>MP: Delhi (+15.4 pp)<br>LP: Chandigarh (+25.5 pp); Jammu and Kashmir (–12.6 pp) | HP: Tamil Nadu (+25.9 pp); Bihar (–12.6 pp) (Figure 3A)<br>MP: Delhi (+31.6 pp) (Figure 3B)<br>LP: Chandigarh (+16.0 pp); Uttarakhand (–10.0 pp) (Figure 3C) | HP: Tamil Nadu (+24.7 pp); Bihar (–12.5 pp)<br>MP: Delhi (+11.7 pp)<br>LP: Uttarakhand (+22.8 pp); Chandigarh (+20.7 pp); Jammu and Kashmir (–14.1 pp) | HP: Tamil Nadu (+10.9 pp); Bihar (–11.7 pp)<br>MP: Delhi (+25.1 pp)<br>LP: Uttarakhand (+16.6 pp) | HP: Tamil Nadu (+17.0 pp); Bihar (–11.6 pp)<br>MP: Delhi (+17.3 pp)<br>LP: Uttarakhand (+16.6 pp) | HP: Tamil Nadu (+14.3 pp); Bihar (–12.3 pp)<br>MP: Delhi (+27.9 pp)<br>LP: Chandigarh (+26.1 pp) |
| **State and UT publications percentage by year**<sup>¶</sup> | Over 2014-2025, the most consistent contributors by publication count in their respective population groups were Tamil Nadu (HP), Delhi and Karnataka (MP), and Chandigarh and Uttarakhand (LP). | Consistent contributors were Tamil Nadu and Uttar Pradesh (HP), Delhi (MP), and Chandigarh and Jammu and Kashmir (LP). | Tamil Nadu and Maharashtra (HP), Delhi and Karnataka (MP), and Chandigarh and Uttarakhand (LP) contributed most steadily. | Most HP states contributed steadily except Bihar and Madhya Pradesh; Delhi (MP) and Uttarakhand (LP) contributed most steadily (Figure 4A-C). | Maharashtra, Tamil Nadu, Uttar Pradesh, and West Bengal (HP), Delhi (MP), and Uttarakhand (LP) contributed most steadily. | Maharashtra, Tamil Nadu, and Uttar Pradesh (HP), Delhi (MP), Chandigarh and Uttarakhand (LP) contributed most steadily. |
| **Publications versus HDI over time**<sup>#</sup> | There is a positive association between HDI and publication volume. Uttar Pradesh in HP and Uttarakhand in LP are outliers, with relatively higher output despite lower HDI. | There is a positive association between HDI and publication volume, with Uttar Pradesh again as an outlier within HP, showing relatively higher output despite lower HDI. | There is a positive association between HDI and publication volume. Uttar Pradesh in HP and Uttarakhand in LP are again outliers, with relatively higher output despite lower HDI. | There is some positive association observed; however, higher HDI states (e.g., Delhi, Maharashtra, Tamil Nadu) account for a large share of output. | There is a positive association between HDI and publication volume. Uttar Pradesh and Uttarakhand are the exceptions (Figure 5). | There is some positive association between HDI and publication volume observed, with Uttar Pradesh as an outlier within HP, with relatively higher output despite lower HDI. |
| **Top three institutions by their publication counts** | Vellore Institute of Technology (Tamil Nadu), SRM Institute of Science and Technology (Tamil Nadu), Chandigarh University (Punjab). | Punjab Agricultural University (Punjab), Indian Agricultural Research Institute (Delhi), International Crops Research Institute for the Semi-Arid Tropics (Telangana). | Vellore Institute of Technology (Tamil Nadu), Chandigarh University (Punjab), SRM Institute of Science and Technology (Tamil Nadu). | University of Delhi (Delhi), Jawaharlal Nehru University (Delhi), Indian Institute of Technology Delhi (Delhi). | University of Delhi (Delhi), University of Petroleum and Energy Studies (Uttarakhand), Vellore Institute of Technology (Tamil Nadu). | All India Institute of Medical Sciences (Delhi), Postgraduate Institute of Medical Education and Research (Chandigarh), Government Medical College (Kerala). |

Note:
\*Unless otherwise stated, counts use the **First author** rule. A state/UT receives 1 count if the first author's first Indian institution is located there. Publications with no India-affiliated first author do not contribute to any count.
\*High population group is defined as states/UTs with an estimated population share of over 5.0% of India's total population; medium population group is defined as states/UTs with an estimated population share between 1.0% and 5.0% (inclusive) of India's total population; and low population group is defined as those with less than 1.0% of the national population. The population estimates were extracted from tables reporting projected total populations of states and UTs from the *Population Projections for India and States 2011-2036* report published by the National Commission on Population under the Ministry of Health and Family Welfare, Government of India [14].
What each row reports:
† Report top five states/UTs by their publication counts.
‡ Report national and state/UT time trends.
§ Report states/UTs where the absolute difference between their share of publications and their share of population is greater than ±10.0 pp in their respective population groups; positive values indicate overrepresentation and negative values indicate underrepresentation.
¶ Describe how each state/UT's share of publications changes over time within its population group (high, medium, low), highlighting states/UTs that contribute consistently.

## Key Findings

1. Publication output overall is highly concentrated in a small set of states/UTs, including Delhi, Tamil Nadu, Maharashtra, and Uttar Pradesh.
2. Most research domains show publication growth from 2014 to 2025.
3. Tamil Nadu and Delhi were persistently overrepresented in the high and medium population groups, respectively, with publication shares more than 10.0 pp higher than their population shares. Bihar was underrepresented in all research domains, with publication shares more than 10.0 pp lower than its population share. Chandigarh and Uttarakhand were frequently overrepresented within the low population group.
4. Tamil Nadu, Uttar Pradesh, Maharashtra, Delhi, Chandigarh and Uttarakhand account for a consistent share of publications over time in their respective population groups, in which first author leadership roles are repeatedly anchored in a few research hubs.
5. Higher HDI states generally publish more, but notable exceptions (e.g., Uttar Pradesh and Uttarakhand) show high output despite lower HDI, indicating spaces where capacity and collaboration may outweigh development levels.
6. In EHR- and AI-related research, most of the top three institutions by publication count were based in Tamil Nadu (EHR: Vellore Institute of Technology and SRM Institute of Science and Technology both from Tamil Nadu, and Chandigarh University from Punjab; AI: Vellore Institute of Technology, Chandigarh University from Punjab, and SRM Institute of Science and Technology). In GWAS, environmental science, and COVID-19-related research, the top three institutions were distributed across a mix of different states and union territories. In development economics, all three top institutions were based in Delhi (University of Delhi, Jawaharlal Nehru University, Indian Institute of Technology Delhi).

## Discussion

Our web application **Indiapub** is designed to help users quickly identify broad patterns in research participation across India and to compare states and UTs on a common visual platform. By making these patterns accessible through interactive displays, this tool can help researchers, trainees, administrators, and policymakers recognize areas of concentration and potential underrepresentation in the national research landscape. Our 2014 to 2025 “Any Topics” snapshot shows that the states/UTs that had a large research participation footprint were among the larger and more populated states and union territory—Tamil Nadu, Maharashtra, Delhi, Uttar Pradesh, and Karnataka. The clustering is also institutional, as the top three publishing institutions were in Tamil Nadu and Delhi, underscoring state and UT-level dominance in research participation and collaboration in India.

The mix of mature and emerging topics allows for a holistic comparison of publication volume at national and state/UT levels, revealing states/UTs with long-established research capacities and those who are pioneers in new research areas. From 2014 to 2025, COVID-19-related research constitutes the largest volume overall, driven by the surge in publications after the outbreak in late 2019. This pattern underscores India’s ability to leverage research capacity and promptly respond to pressing scientific inquiries. AI-related research shows the next highest publication output, followed by environmental science, while EHR, development economics, and GWAS appear as smaller, more specialized areas with comparatively fewer publications.

Comparing each state’s/UT’s share of publications with its share of the national population within its population-size peer group (high, medium, or low) showed that publication output is not proportional to demographic weight. Applying an absolute threshold of 10.0 percentage points between a state’s/UT’s publication share and its population share, we revealed that Tamil Nadu was consistently overrepresented, while Bihar was consistently underrepresented across all six research domains in the high population group. Delhi was overrepresented for all research domains within the medium population group. Chandigarh and Uttarakhand were frequently overrepresented relative to the states and UTs belonging to the low population group, suggesting that concentrated institutional resources supported certain states and UTs with smaller populations (Table 3).

Longitudinal analyses showed that publication counts were driven by population-dense states/UTs such as Tamil Nadu, Uttar Pradesh, Maharashtra, and Delhi. Higher HDI was generally associated with greater publication output. Within the high population group, Uttar Pradesh was a notable outlier: despite a relatively low HDI (ranging from 0.56 to 0.60), it showed large publication output, unlike other low-HDI, high population states such as Bihar (ranging from 0.56 to 0.58) and Rajasthan (ranging from 0.64 to 0.66). These patterns illustrate how **Indiapub** can support evidence-informed planning for research capacity building across states and UTs in India.

To examine whether these publication volume-driven patterns persist when focusing on higher-impact publications, we repeated our case study analyses restricting to publications with at least 50 citations (Table 4). This threshold substantially reduced publication counts across all domains (e.g., from 3,106 to 86 publications in EHR, from 1,431 to 98 in GWAS, and from 48,472 to 1,650 in AI). Several broad patterns were similar to those observed without a citation threshold: Tamil Nadu and Delhi continued to be overrepresented in their respective population groups, and Bihar remained consistently underrepresented in the high population group. However, the top three institutional landscape changed by at least one institution in research domains EHR, AI, development economics, environmental science, and COVID-19. For example, in development economics-related publications, the top three institutions shifted from being exclusively based in Delhi to Malaviya National Institute of Technology Jaipur (Rajasthan) as the leading institution among publications with at least 50 citations, followed by Anna University, Chennai (Tamil Nadu), and Indian Institute of Technology Kharagpur (West Bengal).

**Table 4.**
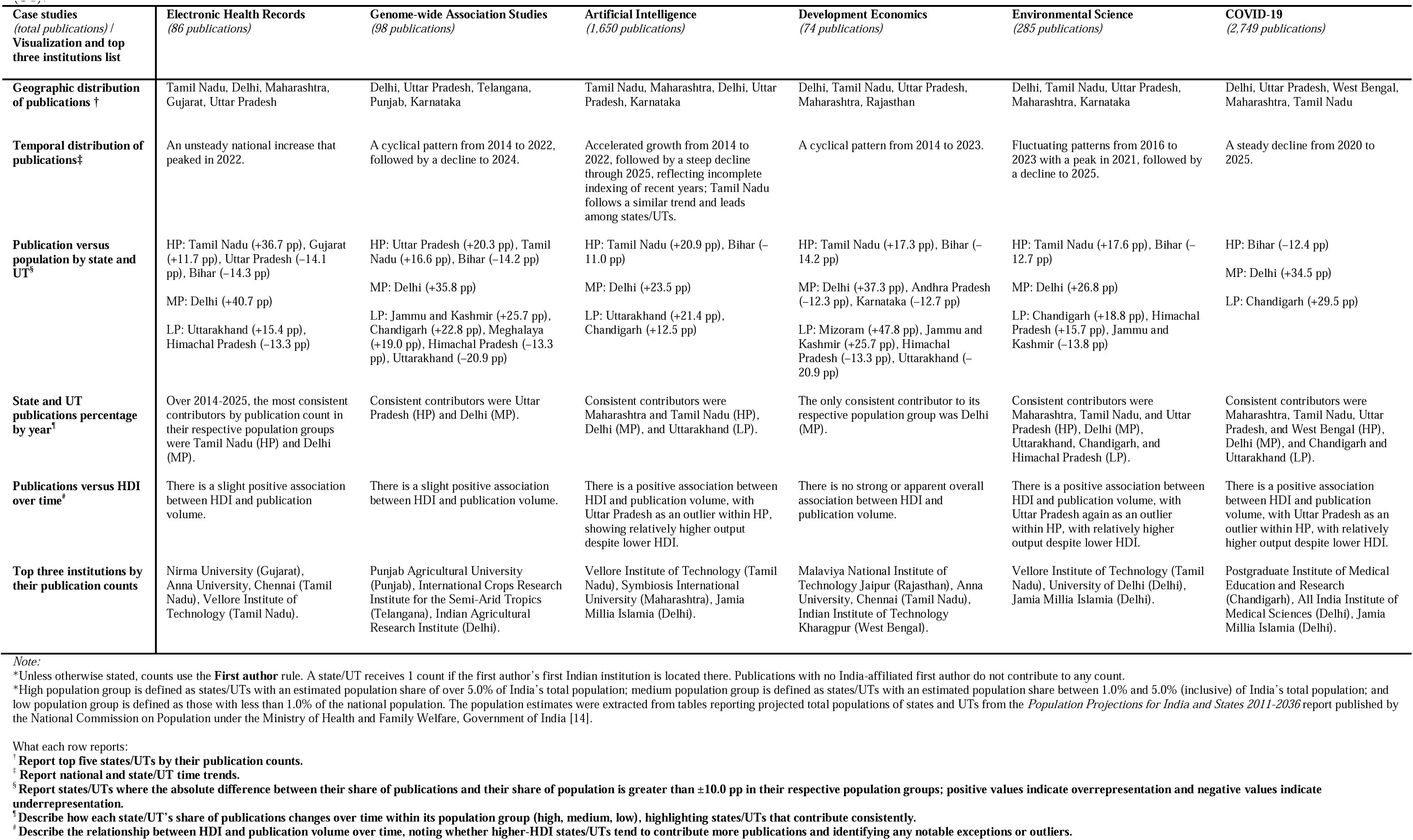
Details of case study findings from visualizations in the **Indiapub** web application covering the topics of electronic health records (EHR), genome-wide association studies (GWAS), artificial intelligence (AI), development economics, environmental science, and COVID-19 across the period 2014-2025. The minimum citations threshold is 50. The results derived from the application are as of March 10, 2026. Abbreviations: High population group (HP), medium population group (MP), low population group (LP), Human Development Index (HDI), National Capital Territory of Delhi (Delhi), percentage points (pp), union territory (UT).

**Figure 1.**
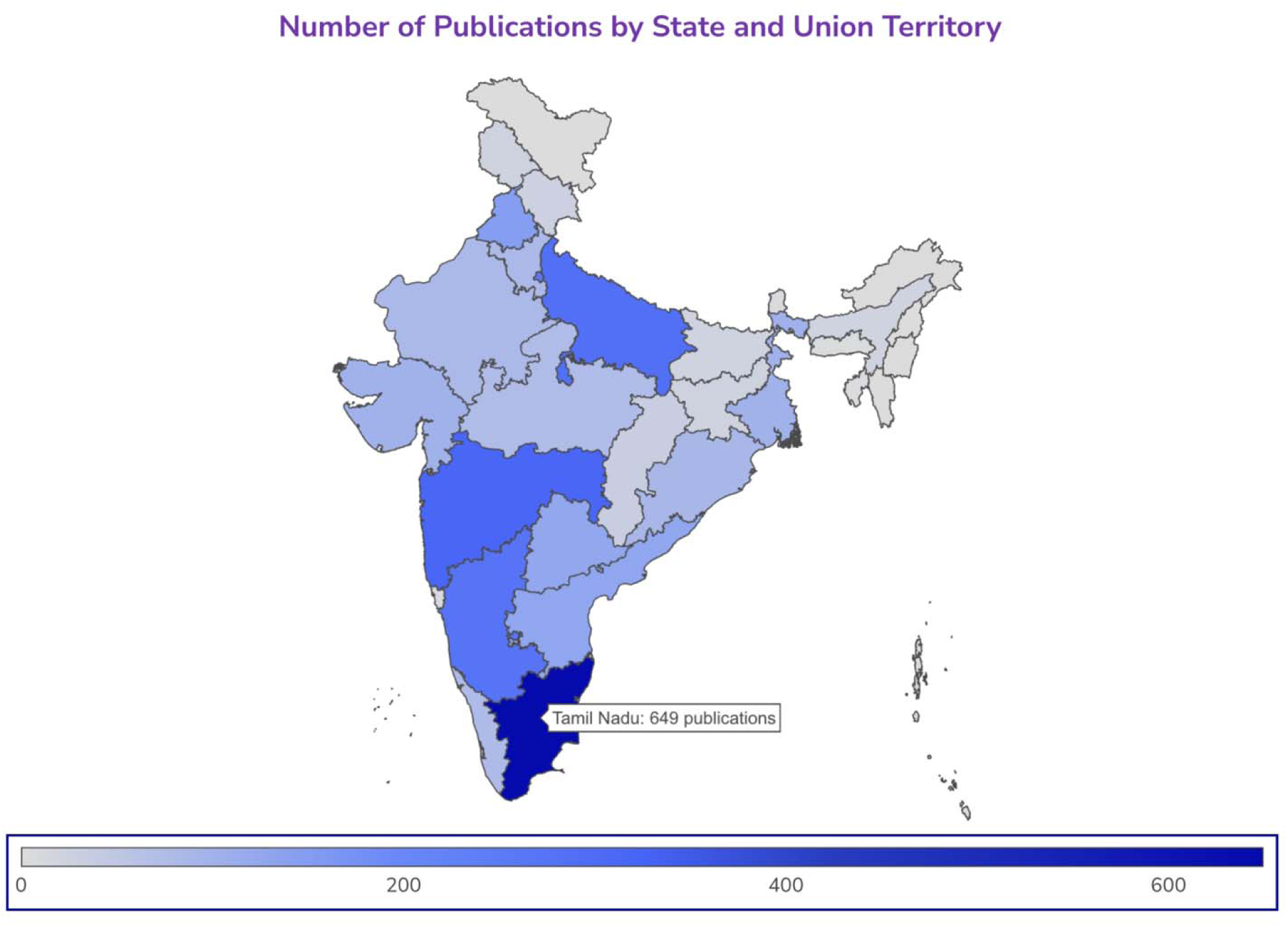
Geographic distribution of publications in the electronic health records research domain from 2014 to 2025. The results derived from the application are as of March 10, 2026.

**Figure 2.**
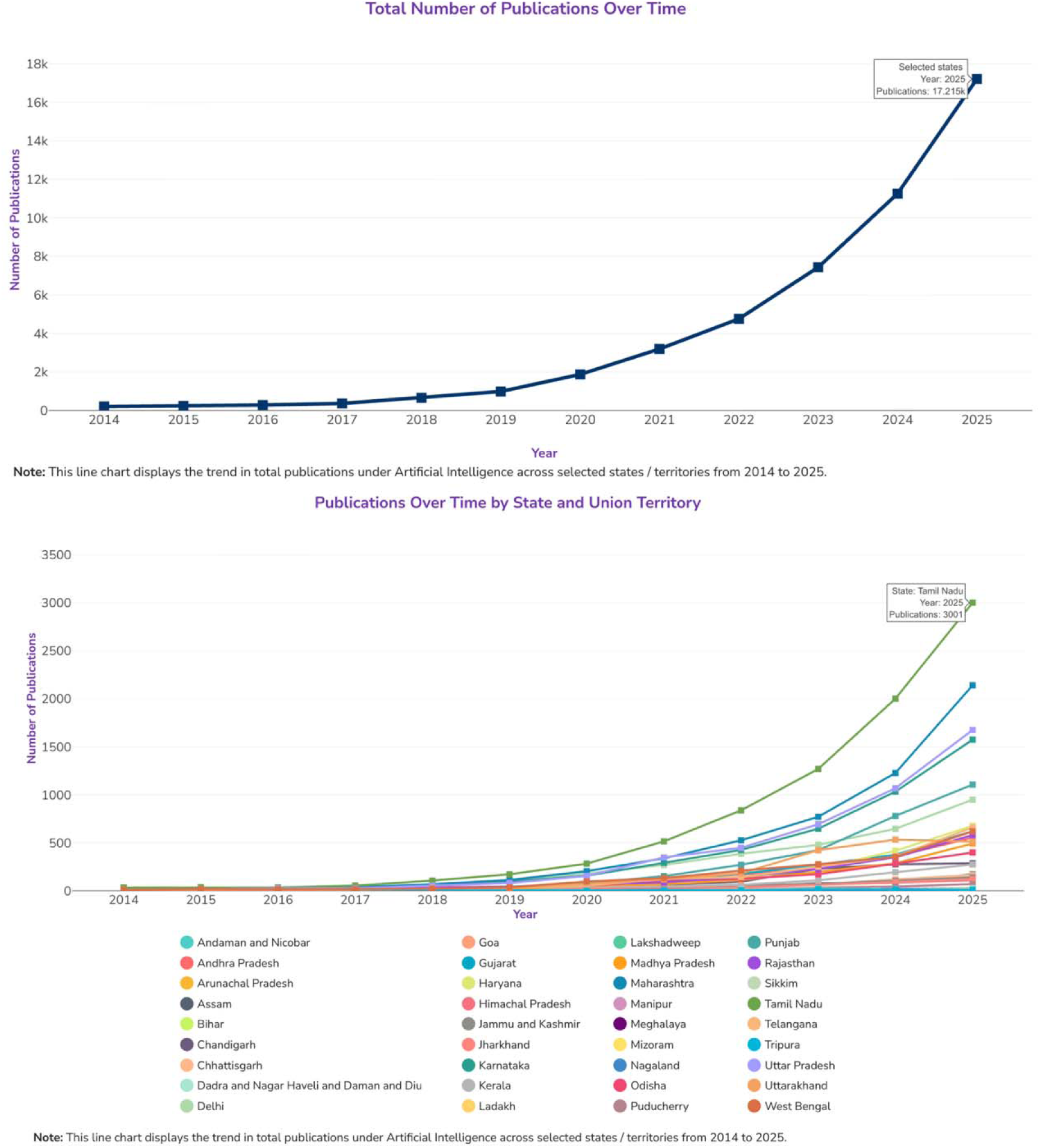
Temporal distribution of publications in the artificial intelligence research domain from 2014 to 2025. Panel A shows the national trend, and Panel B shows the state/UT trends. The results derived from the application are as of March 10, 2026.

**Figure 3.**
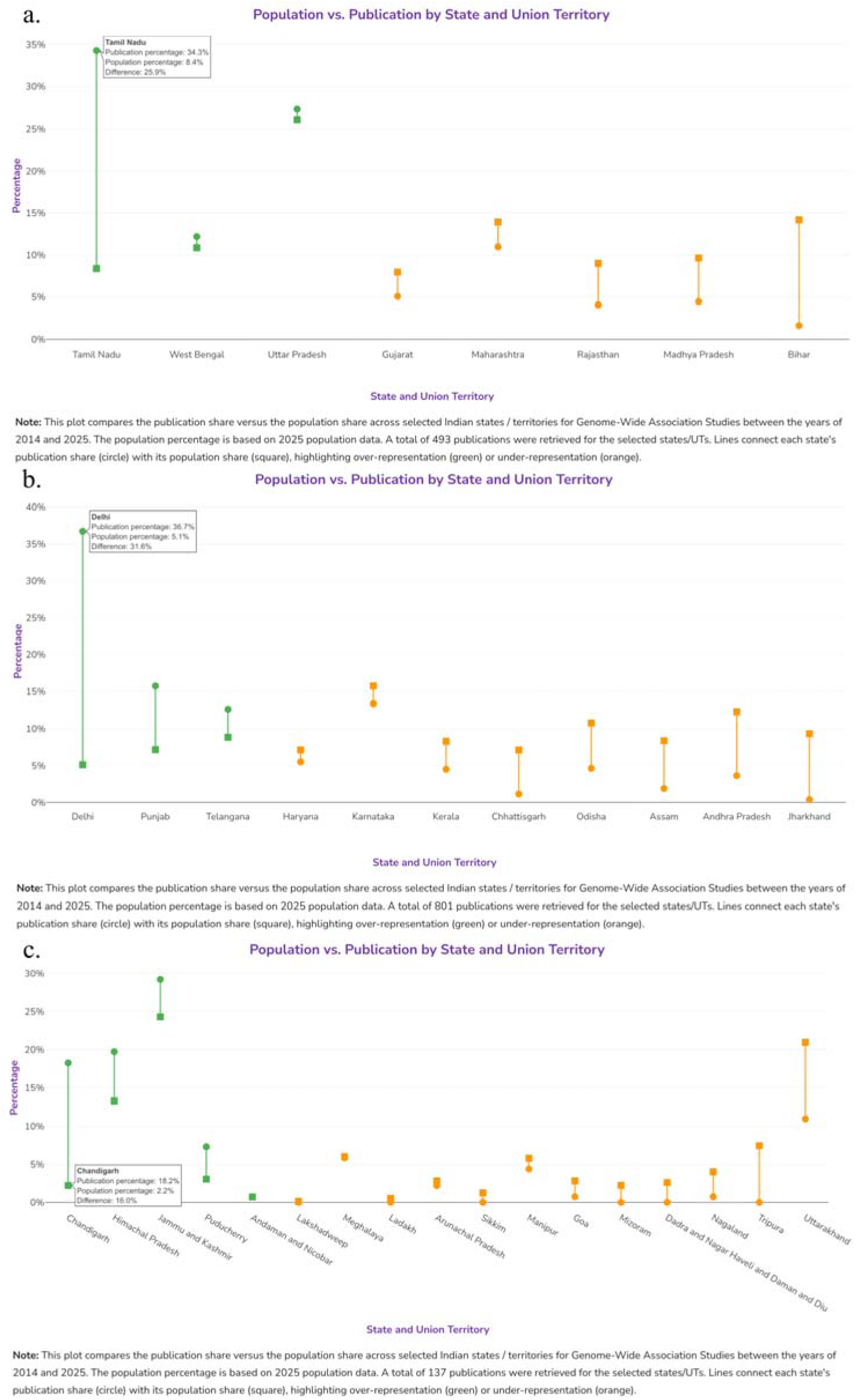
Publication versus population share across high (panel A), medium (panel B), and low (panel C) population groups in the genome-wide association studies research domain from 2014 to 2025. The results derived from the application are as of March 10, 2026.

**Figure 4.**
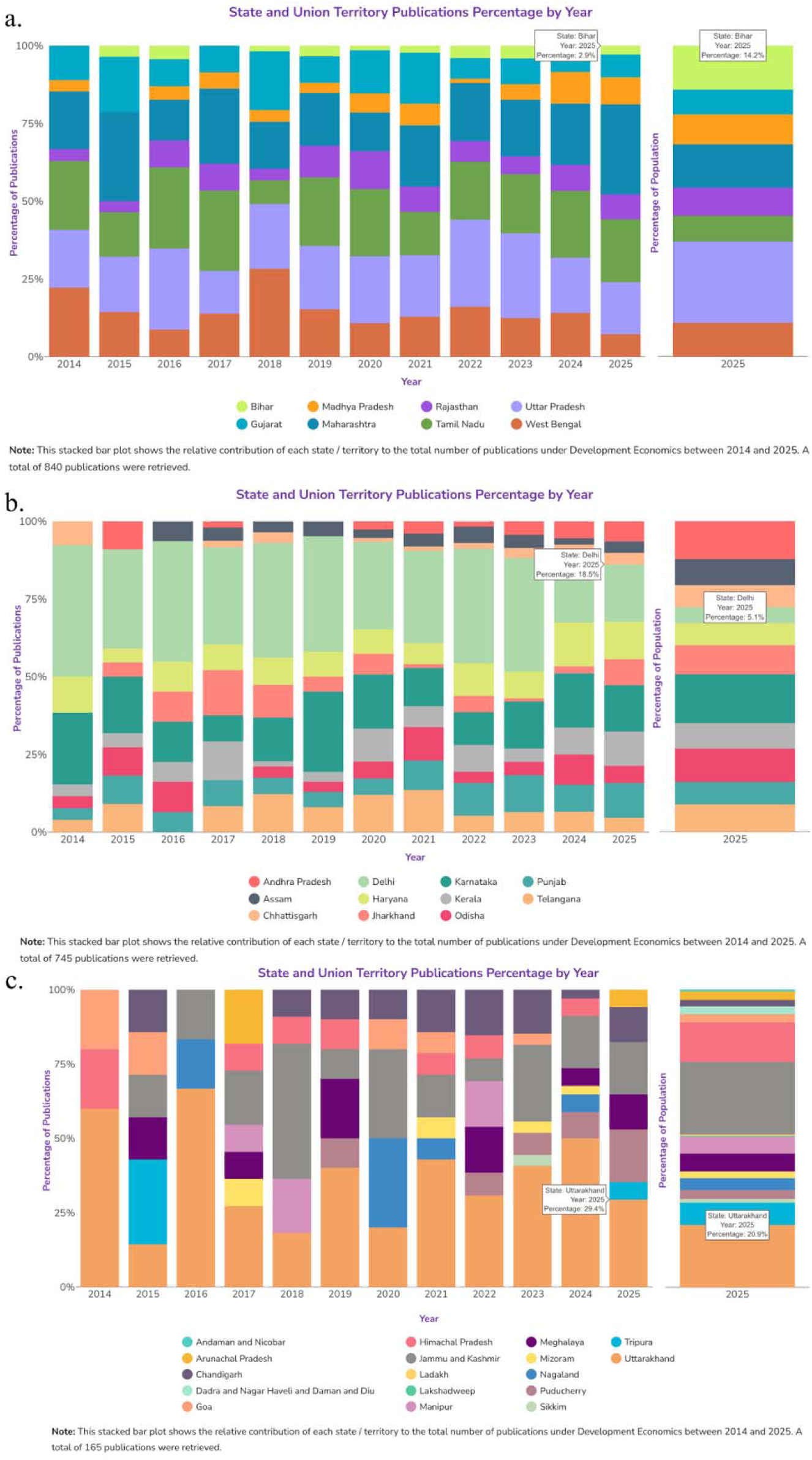
State and union territory publications percentages by year across high (panel A) medium (panel B), and low (panel C) population groups in the development economics research domain from 2014 to 2025. The results derived from the application are as of March 10, 2026.

**Figure 5.**
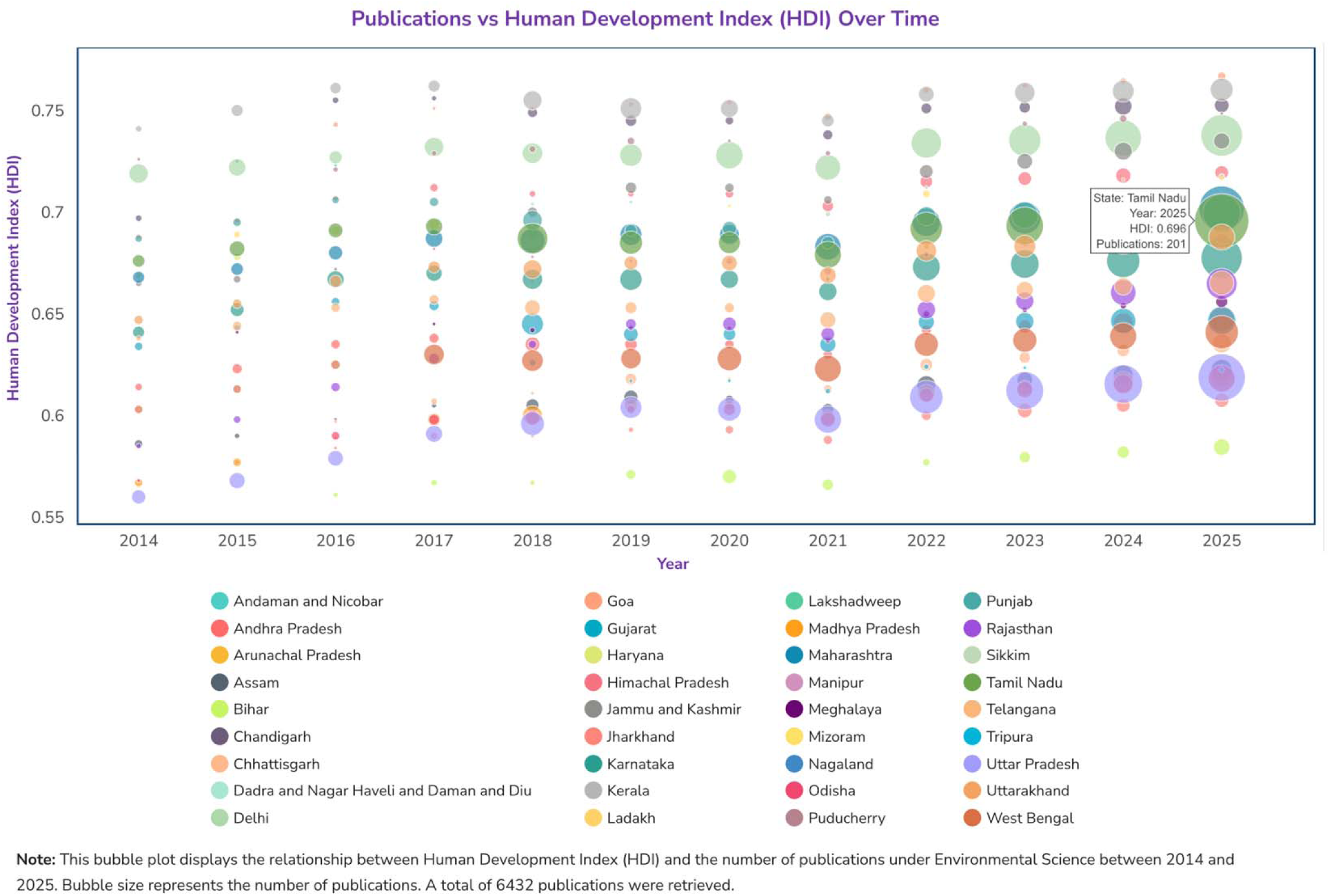
Publications versus Human Development Index over time in the environmental science research domain from 2014 to 2025. The results derived from the application are as of March 10, 2026.

We would like to point out that **Indiapub** is meant to characterize the research capacity of institutions and does not represent the distribution of the studied populations (for studies that involve empirical data on specific populations or geography). State- and UT-level publication counts should be interpreted as measures of institutional research participation and affiliation-based scholarly output, not necessarily as indicators of research focused on that same geography. Researchers based in one state or UT may conduct studies in other regions, use national or international data or no data, or collaborate across multiple institutions and locations. Accordingly, the geographic patterns displayed in **Indiapub** may not correspond directly to the beneficiaries of the research.

Additionally, our analysis relies on OpenAlex-indexed journals and ROR-based affiliations; therefore, publications or affiliations missing from these sources are not captured. Results are descriptive, as we report differences in publication share versus population share without uncertainty quantifications or formal statistical tests. State/UT-level HDI and population share offer only partial insight into structural research capacity in India. Additional factors, such as the number and density of research institutions, university budgets, faculty-to-student ratios, and institution collaborations, are likely influential to publication outcomes. Possible extensions, including incorporating district-level data, co-authorship networks, and funding flows, could provide a more granular view of India’s research ecosystem. Integrating additional bibliographic sources, incorporating uncertainty summaries for comparative plots, and connecting data on social determinants can deepen interpretation of India’s research development and growth.

## Conclusion

As national totals of publication output can obscure large within-country variations, we developed an open-access, interactive web application that allows users to explore publication trends across Indian states and UTs through an automated system of data retrieval, cleaning, aggregation, and visualization. The application serves multiple types of users by making research capacity in India across states/UTs visible through quantifying publication output. For students and early-career researchers, it offers a navigable view of India’s research ecosystem, highlighting active domains and leading institutions for potential collaboration or study. For funders, institutional leaders, and policymakers, it provides an evidence base to guide equitable investment in research infrastructure. Our intuitive visualizations through publication data make it easy to see where research capacity is concentrated, where it is emerging, and where targeted support could accelerate such growth in India. Our platform can be adapted to other countries and topics, which would allow for a reusable template for quantitative enumeration of publication trends.

## Data Availability

The data used in this application are publicly available from the OpenAlex database.

## Code Availability

The code used in this study is available through the Yale School of Public Health Data Science and Data Equity GitHub repository: https://github.com/ysph-dsde/india-publications. Code is updated in the GitHub repository except the “Any Topics.” If you are interested in the “Any Topics” code, please contact the Yiren Hou for the ZIP file. We intend to maintain the **Indiapub** web application and public access to it through at least December 2027, subject to continued server and institutional support.

## Author Contributions

Conceptualization: BM

Supervision: BM

Methodology: BM, YR, KN, YH, JH, EC

Application creation: EC, YR, JH, YH

Writing – original draft: YH, YR

Writing – review & editing: YH, JH, LR, EC, YR, KN, BW, BM

## Acknowledgements

We thank Parikshit Ghosh, PhD, Department of Economics, Delhi School of Economics, University of Delhi, India, for his helpful input in identifying economics-related search keywords for one of the case studies.

